# Meta-analyses on SARS-CoV-2 Viral Titers in Wastewater and Their Correlations to Epidemiological Indicators

**DOI:** 10.1101/2022.02.14.22270937

**Authors:** David Mantilla-Calderon, Kaiyu (Kevin) Huang, Aojie Li, Kaseba Chibwe, Xiaoqian Yu, Yinyin Ye, Lei Liu, Fangqiong Ling

## Abstract

**Background:** Recent applications of wastewater-based epidemiology (WBE) have demonstrated its ability to track the spread and dynamics of COVID-19 at the community level. Despite the growing body of research, quantitative synthesis of SARS-CoV-2 titers in wastewater generated from studies across space and time using diverse methods has not been performed.

**Objective:** The objective of this study is to examine the correlations between SARS-CoV-2 viral titers in wastewater across studies, stratified by key covariates in study methodologies. In addition, we examined the associations of proportions of positive detections (PPD) in wastewater samples and methodological covariates.

**Methods:** We systematically searched the Web of Science for studies published by February 16^th^, 2021, performed a reproducible screen, and employed mixed-effects models to estimate the levels of SARS-CoV-2 viral titers in wastewater samples and their correlations to case prevalence, sampling mode (grab or composite sampling), and the fraction of analysis (FOA, i.e., solids, solid-supernatant mixtures, or supernatants/filtrates)

**Results:** A hundred and one studies were found; twenty studies (1,877 observations) were retained following a reproducible screen. The mean of PPD across all studies was 0.67 (95%-CI, [0.56, 0.79]). The mean titer was 5,244.37 copies/mL (95%-CI, [0; 16,432.65]). The Pearson Correlation coefficients (PCC) between viral titers and case prevalences were 0.28 (95%-CI, [0.01; 0.51) for daily new cases or 0.29 (95%-CI, [-0.15; 0.73]) for cumulative cases. FOA accounted for 12.4% of the variability in PPD, followed by case prevalence (9.3% by daily new cases and 5.9% by cumulative cases) and sampling mode (0.6%). Among observations with positive detections, FOA accounted for 56.0% of the variability in titers, followed by sampling mode (6.9%) and case prevalence (0.9% by daily new cases and 0.8% by cumulative cases). While sampling mode and FOA both significantly correlated with SARS-CoV-2 titers, the magnitudes of increase in PPD associated with FOA were larger. Mixed-effects model treating studies as random effects and case prevalence as fixed effects accounted for over 90% of the variability in SARS-CoV-2 PPD and titers.

**Interpretations:** Positive pooled means and confidence intervals in PCC between SARS-CoV-2 titers and case prevalence indicators provide quantitative evidence reinforcing the value of wastewater-based monitoring of COVID-19. Large heterogeneities among studies in proportions of positive detections, titers, and PCC suggest a strong demand in methods to generate data accounting for cross-study heterogeneities and more detailed metadata reporting. Large variance explained by FOA suggesting FOA as a direction that needs to be prioritized in method standardization. Mixed-effects models accounting for study level variations provide a new perspective to synthesize data from multiple studies.

## 1. INTRODUCTION

Wastewater-based virus monitoring has been shown as a promising tool for tracking disease dynamics in a large population during the ongoing COVID-19 pandemic (Larsen and Wigginton, 2020). It has been reported that 39 to 65% of infected individuals may excrete viral particles through urine and feces (Chen, Chen, et al., 2020; Xiao, Sun, et al., 2020; Guo, Tao, et al., 2021), thus allowing wastewater-based detection. Wastewater-based epidemiology (WBE) has the potential to circumvent biases caused by varied accesses to individual-based testing, presence of asymptomatic cases, and social stigma (Murakami, Hata, et al., 2020). Moreover, WBE has been applied in environmental monitoring of polioviruses, effectively detecting new variants and preventing disease resurgence (Hovi, Shulman, et al., 2012). This past experience suggests the long-term benefits in the development and refinement of WBE as a public health monitoring technology.

While the number of WBE studies continues to grow, it demands attention to seek generalizable relationships across studies and account for study-to-study variations. For example, though WBE studies focusing on SARS-CoV-2 viruses are all conducted during the pandemic, not all wastewater samples tested positive for SARS-CoV-2 by qPCR/dPCR (Medema, Heijnen, et al., 2020; Gonzalez, Curtis, et al., 2020; Gonçalves, Koritnik, et al., 2021; Graham, Loeb, et al., 2020; Ahmed, Angel, et al., 2020; Baldovin, Amoruso, et al., 2021; Hata, Hara-Yamamura, et al., 2021; Kitamura, Sadamasu, et al., 2021; Nemudryi, Nemudraia, et al., 2020; Saguti, Magnil, et al., 2021; Sherchan, Shahin, et al., 2020; Trottier, Darques, et al., 2020; Randazzo, Truchado, et al., 2020). In addition, while positive correlations between SARS-CoV-2 wastewater-based measurements and COVID-19 cases have been described (Medema, Heijnen, et al., 2020; Gonzalez, Curtis, et al., 2020; Graham, Loeb, et al., 2020; D’Aoust, Mercier, et al., 2021; D’Aoust, Graber, et al., 2021; Peccia, Zulli, et al., 2020), the strength of the correlations may vary among studies. To fully untap the potentials of WBE, research synthesis efforts are needed to quantify detection rates of SARS-CoV-2 in wastewater, its titers, and their correlations to epidemiological indicators.

Meta-analysis provides an objective, quantitative, and powerful way to synthesize findings across studies (Gurevitch, Koricheva, et al., 2018). This analysis methodology treats individual studies as members of a population of studies that all provide information on a given effect instead of drawing conclusions on exemplary studies that have shown strong positive effects (Murad and Montori, 2013). Leveraging a large sample size, meta-analysis can help move the narrative beyond statistical significance, and draw attention to the magnitude, direction, and variance in effects (Gurevitch, Koricheva, et al., 2018). Furthermore, the meta-analytic approach allows us to quantitatively examine heterogeneity among study results, thus motivating the generation of new hypotheses (Linden and Hönekopp, 2021). Meta-analysis is considered beneficial in medicine (Murad and Montori, 2013), social science (Card, 2015), ecology and evolution (Arnqvist and Wooster, 1995), among other fields. Because of the interdisciplinary nature of WBE, a meta-analysis about the booming literature providing pooled effect sizes and heterogeneity can provide quantitative evidence to help justify research to refine and advance the technology by interested researchers from multiple fields.

Here, we employed a meta-analytic methodology to synthesize wastewater-based SARS-CoV-2 viral titer data published by February 16th, 2021, approximately a year after the beginning of the pandemic. Following a reproducible pipeline (PRISMA guideline, Page, McKenzie, et al., 2021), we synthesized results from 1,877 observations in 20 studies. We asked four fundamental questions; 1) what is the pooled proportion of positive detection of SARS-CoV-2 from wastewater samples; 2) what are the titers of SARS-CoV-2 viruses in wastewater collectively and when subgrouped by key methodological variables; 3) what are the overall strengths of correlation between positive detection or titers of SARS-CoV-2 in wastewater to epidemiological indicators (active and cumulative cases); 4) how much of the variation in SARS-CoV-2 viral titers can be explained by COVID-19 cases alone? Mixed effects models were employed to examine correlation between SARS-CoV-2 viral titers and detection in wastewater while accounting for study-level variations.

## 2. METHODS

This systematic review and subsequent meta-analysis have been conducted according to the PRISMA guidelines (Page, McKenzie, et al., 2021). A PRISMA checklist is presented in Table S1.

### 2.1 Data sources

We searched Web of Science (WoS) for any publications analyzing untreated wastewater for SARS-CoV-2 viruses on February 16^th^, 2021. Studies were retrieved with the search terms: TS = (SARS-CoV-2 AND (wastewater OR sewage)) from the WoS core collection. The following search conditions were applied: i) language was restricted to English; ii) time span was set to “All Years”; iii) records in the Science Citation Index Expanded (SCI-Expanded) were included; and iv) document type was set to “article”, “early access” or “letter” to retain original research (i.e., “reviews” or “editorial materials” were not selected).

### 2.2 Study selection and eligibility criteria

Upon study retrieval from Web of Science, any duplication of records was screened by titles and authors. No duplication was found. Next, full text records were scanned to assess for eligibility. All studies that reported nucleic-acid detections of SARS-CoV-2 from wastewater systems and associated epidemiological indicators were included. Specifically, inclusion criteria were applied: 1) original qPCR data in terms of SARS-CoV-2 measurements in wastewater were reported, and data were reported as quantification cycle, copy numbers/volume, genome equivalents/volume, or genome equivalents/weight of sample; 2) sampling locations were identified as wastewater treatment plants (WWTP), sewage collection networks, lift stations, manholes or septic tanks; 3) SARS-CoV-2 case incidences/prevalences are reported for the associated locations during the sampling times. Rationales for each inclusion criterion are provided in Table 1. The study eligibility was performed by David Mantilla-Calderon (DMC), Kevin Huang (KH), Aoji Li (AL) and Fangqiong Ling (FL). In case of uncertainties, these were discussed and resolved by consensus.

**Table 1.**
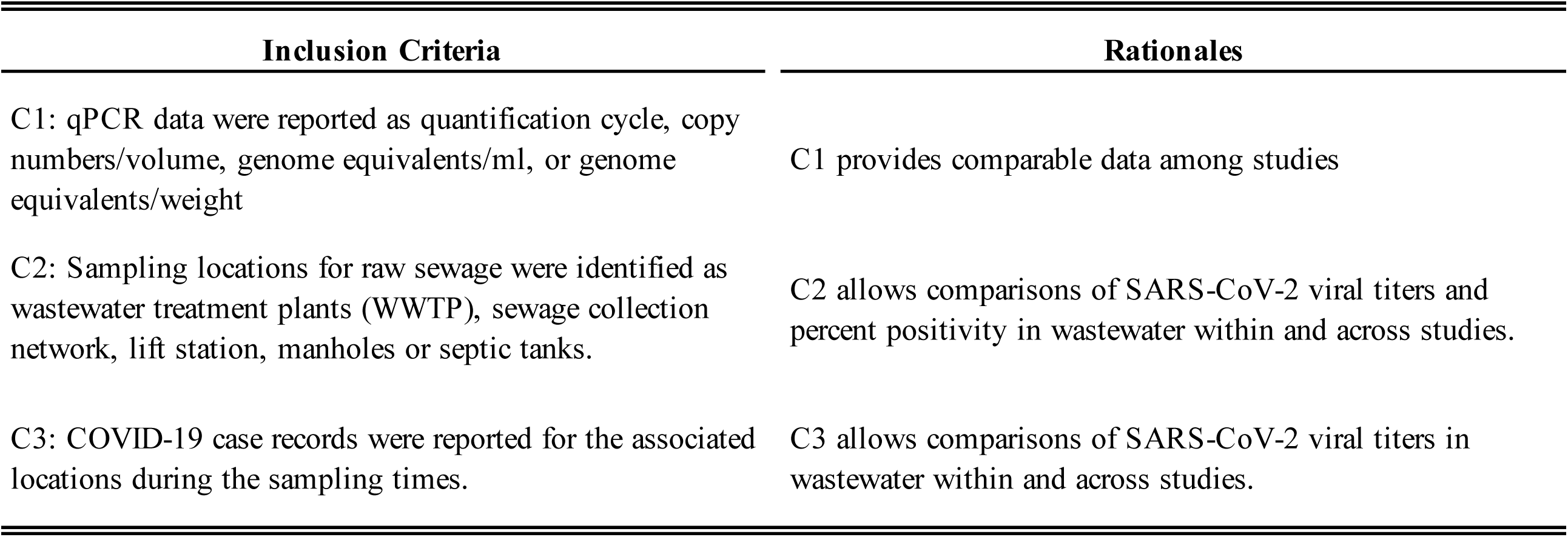
Inclusion criteria for eligibility of studies

### 2.3 Data extraction

SARS-CoV-2 measurements in wastewater were extracted from the full texts or supplementary materials. WebPlotDigitalizer version 4.4 (https://automeris.io/WebPlotDigitizer) was used to retrieve the information using digitized versions of figures. The qPCR/dPCR measurements themselves and the units (i.e., quantitative cycles [Cq], or titers) were recorded. In addition, metadata about a sample was retrieved, including study information, sample environment, and assay information. Study information included author, title, and the year of publication. Sample environment included the following: i) the geographical location (i.e., country and city where the study was performed); ii) sampling location within a wastewater system (i.e., samples were taken from the sewage collection systems, at the wastewater treatment plant after screens and before sedimentation, or at the primary sedimentation tank); iii) sample processing prior to viral concentration (whether a sample was filtrated, centrifuged or left untreated); iv) viral concentration method; v) the associated COVID case incidence or prevalence as provided in the publication; vi) serviced population as provided in the publication, and vii) the date of collection of each wastewater sample. Last but not the least, we extracted assay information, including the choice of sampling techniques (i.e., grab or composite sampling), qPCR gene markers, and primer sets.

### 2.4 Data extraction and summary measures

Upon data retrieval, SARS-CoV-2 measurements, sample environment, assay information, COVID-19 case prevalence were recorded and converted to consistent unit across studies (2.4.1-2.4.3). The proportion of positive detections was calculated (2.4.4). Annotated data will be made available on github.com/linglab-washu/wbe-metaanalysis at the time of publication.

#### 2.4.1 SARS-CoV-2 measurements

SARS-CoV-2 measurements in wastewater were retrieved in terms of copy numbers per mL, genome equivalents (ge) per mL or copy numbers per gram, according to the way measurements were described in the Methods section of each study. Samples that were analyzed using multiple SARS-CoV-2 markers were recorded as multiple entries and denoted under the same sample ID (e.g., if a sample was analyzed for N1, N2 and N3 markers, all measurements would be included in the analysis). In the case that a study reported a range of concentrations for a particular sample (e.g., 1 to 10 copies/mL), the midrange was recorded (e.g., 5 copies/mL). For studies reporting the average and standard deviations of multiple technical replicates for a wastewater sample, only the average was recorded. All eligible studies were included in the systematic review (3.1). Studies providing SARS-CoV-2 measurements as titers or Cq were included in the analysis of positive detections (3.2, 3.4-3.7), and studies that provided titers (copies per unit volume) were included in the analysis on titers (3.3 - 3.7).

#### 2.4.2 Sample environment and assay information

Sample environment and assay information were usually described in free texts and hence manual curation was performed. Some sample environment and assay information had a higher level of semantic consistency (e.g., grab vs. composite samples), while others were described in more varied texts. In the case that varied descriptions had suggested similar meanings, we annotated using consistent texts reflecting the common meanings to facilitate synthesis across studies.

##### Sampling location

Specifically, the sampling locations were annotated as i) “WWTP” if a sample was described as taken after primary screens and before the primary sedimentation tanks, as ii) “municipal sewage network” if a sample was taken from manholes, septic tanks, or lift stations, and as iii) “in premise”, if the sample was described as taken from the private sewage infrastructure of a facility such as a hospital or dormitory.

##### Fractions

Wastewater and sludge samples are included in the review. Wastewater refers to samples collected from the sewage network or the WWTP, consisting predominantly of a liquid phase and to a lesser degree, a solid phase. The term sludge specifically refers to the slurry of solids and liquid mixture collected from a primary clarifier. Liquid and solid phases of samples can be separated/fractionated by laboratory methods. Fractions resulting from sample separation processes were recorded and annotated. Specifically, a fraction was annotated as “Solids” when it consisted of primary solids from a gravity thickener, or solids collected from wastewater by in-laboratory sedimentation (e.g., pellets recovered from centrifugation of the wastewater sample at > 2000 g). A Fraction was annotated as “Supernatant/filtrate” if the sample had been pre-filtered at 0.22 to 0.7 µm or centrifuged at 2.000-10.000 g prior to viral concentration. A fraction was annotated as “Mixture of supernatant and suspended solids” when a sample of raw unprocessed wastewater was directly used for viral concentration.

##### Viral concentration

A fraction might be subjected to a subsequent viral concentration step. Typically, viral concentrations methods were applied to mixture of supernatant and suspended solids and supernatant/filtrate fractions. Solid fractions were not typically subjected to a viral concentration step. Viral concentration methods were recorded as described in the original publication.

##### Gene targets

Several SARS-CoV-2 qPCR gene targets were identified during the study screening process and categorized in the marker variable. Targeted regions included ORF1ab, Nucleoprotein (N), Spike protein (S), Envelop protein (E), Membrane protein (M) and the RNA-dependent RNA-polymerase (RdRp) gene. The specific primer set that was used to target the marker gene was recorded under the primer variable using the notation Author_Marker. Two abbreviations were used in the field author, CDC, referring to the Center of Disease Control, USA (e.g., CDC_N1) and NIID, referring to the National Institute of Infectious Disease, Japan (e.g., NIID_N).

In the cases that combinations of multiple primers were simultaneously used in a qPCR assay, the value for primer for this specific sample was recorded by listing the primer sets spaced by an underscore sign (e.g., CDC_N1_N2). In some instances, a study may analyze multiple markers but report as one genome equivalent. A marker would be recorded if it was specified in the study which marker was used to calculate the genome equivalent; alternatively, the marker recorded for the sample would include all the markers used in the study separated by a comma (e.g., CDC_N1, CDC_N2).

#### 2.4.3 COVID-19 case prevalence

Daily new cases, cumulative cases, and active cases as reported in the papers were retrieved and included in this analysis, with exceptions when i) a sample was collected before epidemiological reporting by local health authorities was available or ii) a sample was collected from sewage lines in buildings in which the incidence of SARS-CoV-2 may significantly differ from city/municipality case reports. Sample entries where consistent case prevalence data were not found in the study, were excluded from the analysis. Case data were recorded in the way that was reported in the study. For samples retrieved from WWTPs that extend over multiple municipalities, case incidence was estimated by computing the average of the case incidence (normalized by population size) of the municipalities contributing to the sewage of that specific WWTP.

Epidemiological data reported as “cumulative cases” are denoted as “cumulative cases”. Cases reported as “daily cases”, “new cases”, “positive daily test”, “new positive daily test”, or “seven-day average cases” were denoted as “daily new cases”. “Hospital admissions” and “hospitalized patients” were denoted as “hospitalized patients”. All case counts were converted to prevalence, i.e., patients per 100,000 inhabitants to allow synthesis across studies.

#### 2.4.4. Proportions of positive detection

Subgroups were defined by different aggregating variables such as study ID, sample collection method, fraction type etc. The proportion of positive detection was defined as the ratio of samples showing positive test results for SARS-CoV-2 in a subgroup, over the total number of samples analyzed in each subgroup.

### 2.5 Forest plot generation

Forest plots were generated using the “dmetar” package for R (Harrer, Cuijpers, et al., 2019) using a random-effects model. A random-effects meta-analysis model assumes the observed average SARS-CoV-2 titers can vary across studies because of real differences in the viruses present in the systems as well as sampling variability (chance). Thus, even if all studies had an infinitely large sample size, the observed study effects would still vary because of the real differences in the sewershed’s effects on viral titers. Such heterogeneity in average viral titers can be caused by differences in study populations (such as local COVID-19 case prevalence), the wastewater system effects on dilution or decay, the methodological differences, and other factors.

Weight of each study in the forest plot was calculated as

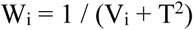

where W_i_ denotes the weight of study_i_, V is the variance and T^2^ (tau) is the variance of each distribution with respect to the grand mean estimated using the Sidik-Jonkman estimator (Sidik and Jonkman, 2005). More details on the calculations can be found at Borenstein et al. (Borenstein, Hedges, et al., 2010). Measurements of SARS-CoV-2 titers were transformed to log copies per mL of sample to allow synthesis across studies and aid with visualization. Observations with SARS-CoV-2 measurements equal to zero marker copies per mL were removed.

### 2.6 General linear mixed effects model (GLMM)

General linear mixed effects models were built to examine the epidemiological indicators (cumulative cases or daily new cases) as sources of fixed effects and study as a source of random effects on SARS-CoV-2 measurements in wastewater. A binomial GLMM was used to model the positive detections among all observations. A linear mixed effects model (Gaussian) was used to fit the log-transformed titers using observations where positive detections were made. GLMMs were built in the R package “lme4” (Bates, Mächler, et al., 2014). Studies were treated as sources of random effects on intercepts. Fixed-effects models were built using the same link functions to examine the significance of random effects and assess the overall fits from fixed effects. Fitting of fixed-effects models were performed using the “glm” and “lm” functions in R stats package (R Core Team, 2013). Akaike information criterion (AIC), Bayesian Information Criterion (BIC) and log likelihoods were reported for model selection. The Nakagwa’s R-squared definitions were used to compute marginal and conditional r-squared values using the R package “MuMIn” (Barton, 2009).The Studies that reported daily new cases and cumulative cases were examined separately. Studies reporting solids were excluded due to lack of replicates after being subset into studies reporting daily new cases or cumulative cases (details can be found in Figure S1).

## 3. RESULTS

### 3.1 Systematic review

Our search identified 101 unique titles and abstracts; after screening (Figure 1, Table S2), 20 papers were included in this review. These studies reported SARS-CoV-2 measurements in wastewater in terms of titers or Ct and provided epidemiological indicators. A total of 1877 observations were recorded. Figure 1 depicts details of the search.

**Figure 1.**
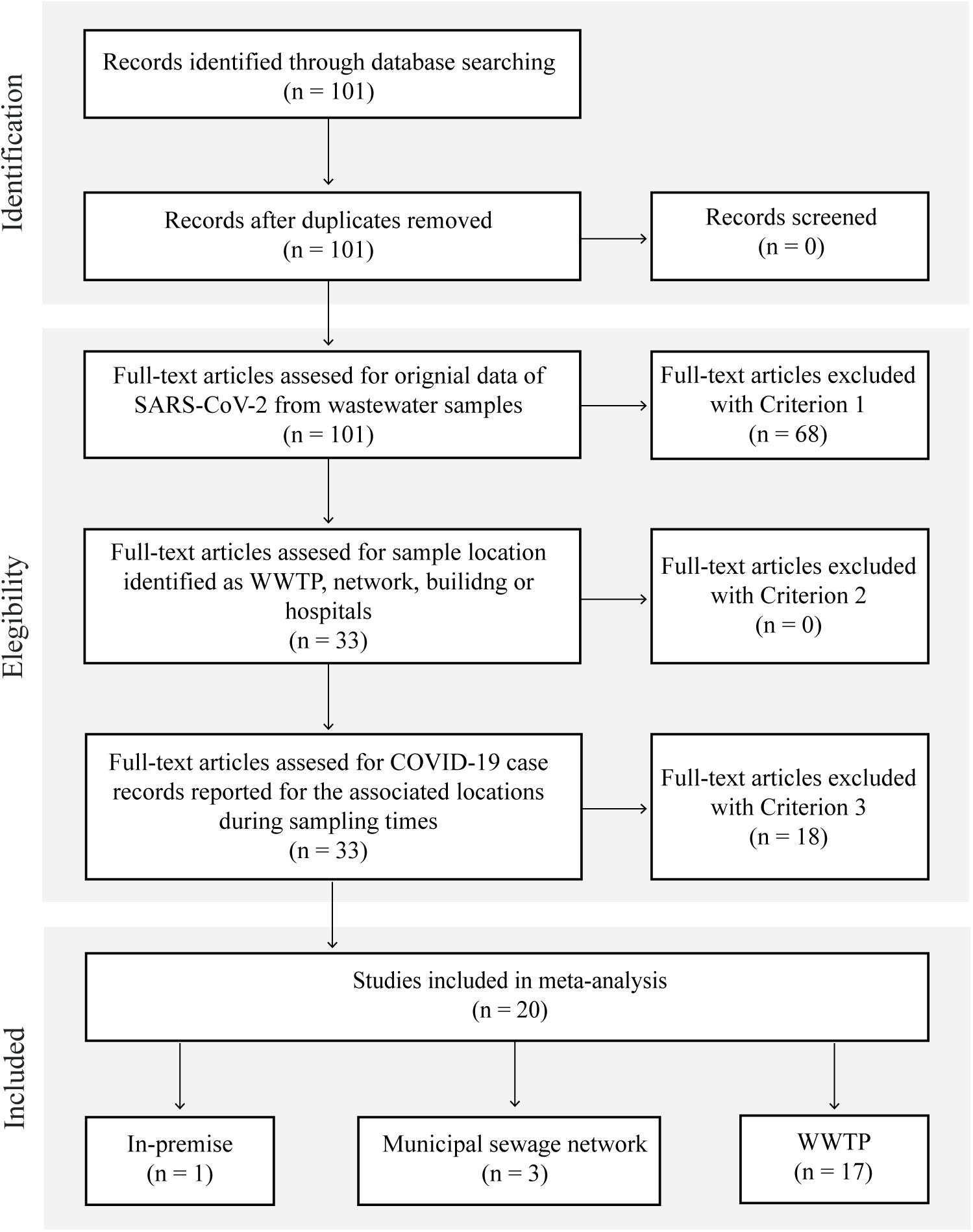
Study selection flow diagram. The three criteria used in the screening for eligibility are: Criterion 1, original data of SARS-CoV-2 from wastewater samples were provided in terms quantification cycle (Cq), copy numbers per unit of volume or weight, or genome equivalents per unit of volume or weight; Criterion 2, sampling location were reported as WWTPs, sewage collection networks, buildings, or hospitals; Criterion 3, COVID-19 case counts of the corresponding times and areas were reported in the study with a clear data source. Reports were found to be primarily sampling at WWTP (17 studies) and less often at municipal sewage network (3 studies) and in-premise (1 study); in-premise sampling location was carried out at a hospital.

Table 2 describes basic characteristics of the included resources. Eighteen studies reported quantitative measurements for SARS-CoV-2 as titers (Medema, Heijnen, et al., 2020; Peccia, Zulli, et al., 2020; Kumar, Patel, et al., 2020; Gonzalez, Curtis, et al., 2020; Graham, Loeb, et al., 2020; D’Aoust, Mercier, et al., 2021; Hata, Hara-Yamamura, et al., 2021; Kitamura, Sadamasu, et al., 2021; Nemudryi, Nemudraia, et al., 2020; Saguti, Magnil, et al., 2021; Sherchan, Shahin, et al., 2020; Trottier, Darques, et al., 2020; Randazzo, Truchado, et al., 2020; Randazzo, Cuevas-Ferrando, et al., 2020; Ahmed, Bertsch, Bivins, et al., 2020; Miyani, Fonoll, et al., 2020; Westhaus, Weber, et al., 2021; Haramoto, Malla, et al., 2020), while two studies reported Ct values (Baldovin, Amoruso, et al., 2021; Gonçalves, Koritnik, et al., 2021). Among the 18 quantitative studies, seventeen reported marker copies or genome equivalent per mL (Medema, Heijnen, et al., 2020; Peccia, Zulli, et al., 2020; Kumar, Patel, et al., 2020; Gonzalez, Curtis, et al., 2020; D’Aoust, Mercier, et al., 2021; Hata, Hara-Yamamura, et al., 2021; Kitamura, Sadamasu, et al., 2021; Nemudryi, Nemudraia, et al., 2020; Saguti, Magnil, et al., 2021; Sherchan, Shahin, et al., 2020; Trottier, Darques, et al., 2020; Randazzo, Truchado, et al., 2020; Randazzo, Cuevas-Ferrando, et al., 2020; Ahmed, Bertsch, Bivins, et al., 2020; Miyani, Fonoll, et al., 2020; Westhaus, Weber, et al., 2021; Haramoto, Malla, et al., 2020), and one study reported marker copies per gram of biomass (Graham, Loeb, et al., 2020). Epidemiological indicators were reported as daily cases in nine studies, ranging from 0.6 per 100,000 inhabitants to 117 per 100,000 inhabitants (Graham, Loeb, et al., 2020; D’Aoust, Mercier, et al., 2021; Baldovin, Amoruso, et al., 2021; Hata, Hara-Yamamura, et al., 2021; Nemudryi, Nemudraia, et al., 2020; Sherchan, Shahin, et al., 2020; Trottier, Darques, et al., 2020; Miyani, Fonoll, et al., 2020; Haramoto, Malla, et al., 2020), cumulative cases were reported in ten studies ranging from 1.6 per 100,000 inhabitants to 808.2 per 100,000 inhabitants (Medema, Heijnen, et al., 2020; Kumar, Patel, et al., 2020; Gonzalez, Curtis, et al., 2020; Hata, Hara-Yamamura, et al., 2021; Kitamura, Sadamasu, et al., 2021; Sherchan, Shahin, et al., 2020; Randazzo, Truchado, et al., 2020; Ahmed, Bertsch, Bivins, et al., 2020; Westhaus, Weber, et al., 2021; Haramoto, Malla, et al., 2020), active cases were reported in four studies (D’Aoust, Mercier, et al., 2021; Gonçalves, Koritnik, et al., 2021; Randazzo, Cuevas-Ferrando, et al., 2020; Westhaus, Weber, et al., 2021) and hospitalized cases in two studies (Baldovin, Amoruso, et al., 2021; Saguti, Magnil, et al., 2021). Among these studies, two studies reported both daily and cumulative cases (Sherchan, Shahin, et al., 2020; Haramoto, Malla, et al., 2020), one study reported both daily and active cases (D’Aoust, Mercier, et al., 2021) and one reported both cumulative cases and hospitalized cases (Baldovin, Amoruso, et al., 2021). Cumulative COVID-19 cases were the most frequently reported, followed by daily, active, and hospitalized cases. SARS-CoV-2 was detected in all studies, irrespective of case prevalence levels, albeit at varying proportions of positive detections.

**Table 2.**
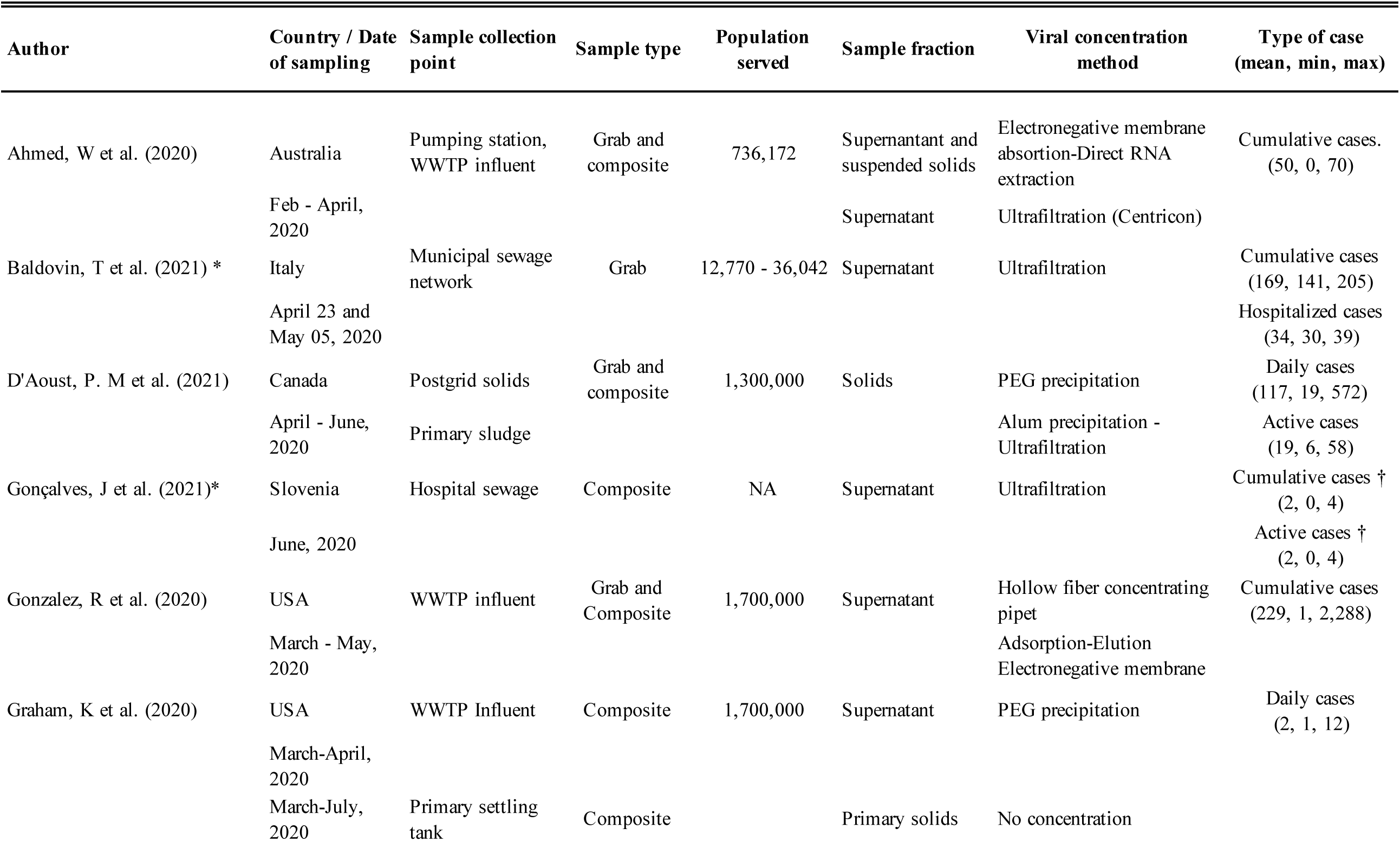
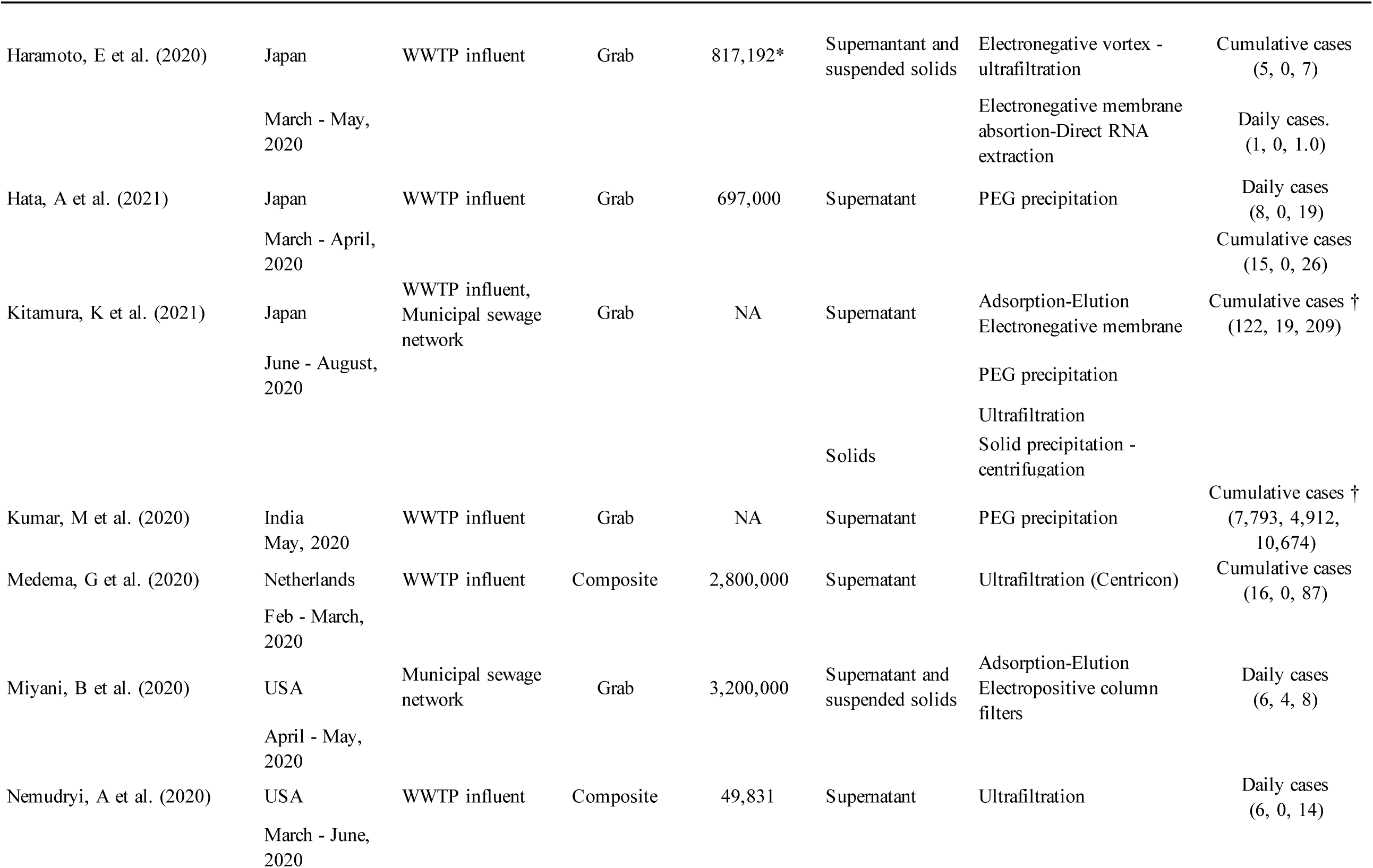
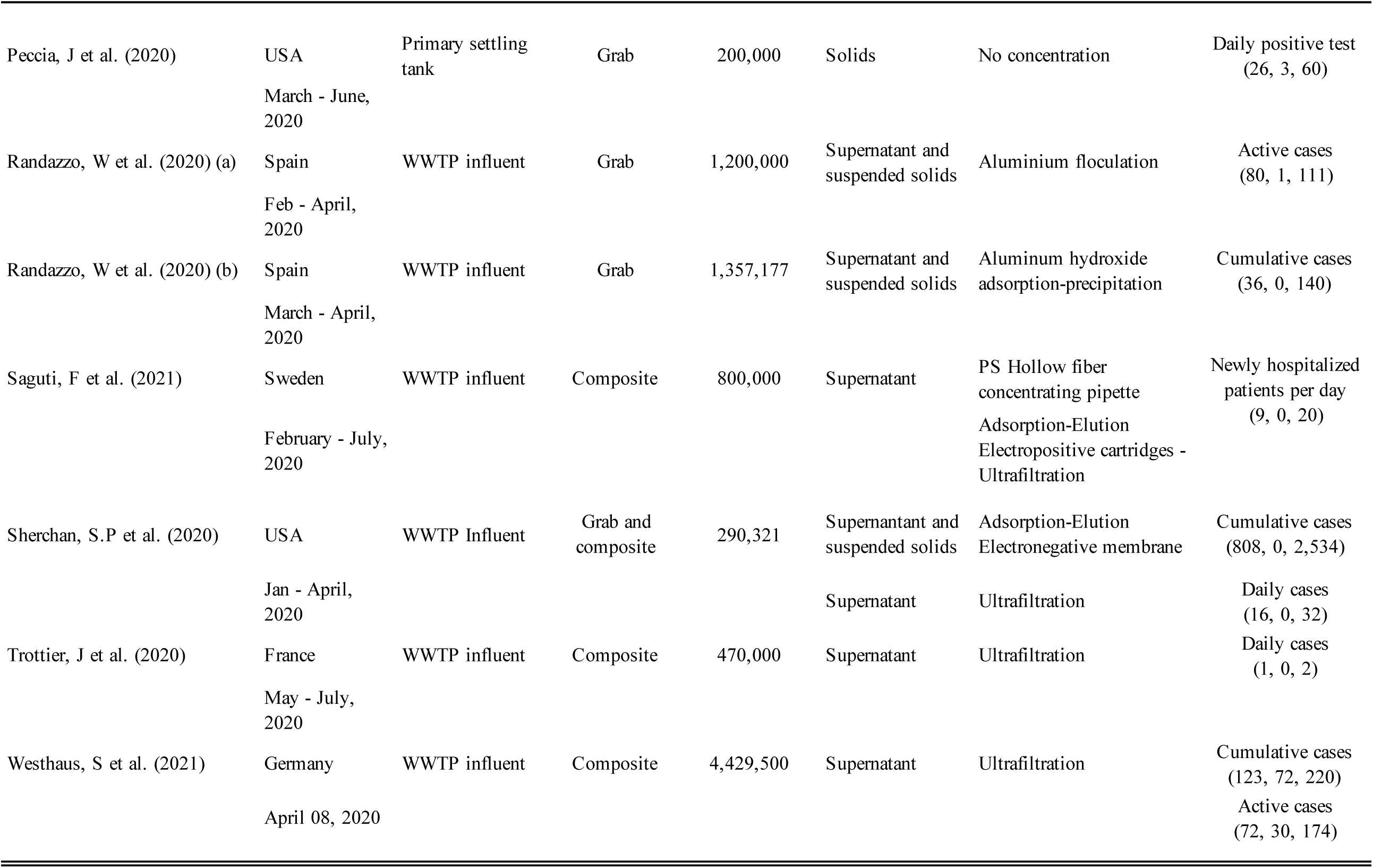
General features of studies included. COVID-19 cases are reported per 100,000 inhabitants unless otherwise stated. COVID-19 cases are rounded to the nearest unit. * Semi-quantitative studies. † cases not normalized by 100,000 inhabitants

Correlations between COVID-19 cases and wastewater SARS-CoV-2 titers were reported in six studies. This is confirmed by our analysis. We performed linear regression on each dataset. Six out of eighteen studies detected significant linear correlations between SARS-CoV-2 titers and the respective epidemiological indicators in the study (*p-value* < 0.05, Table 3, Figure S2-S4). All these six studies were conducted at WWTPs, amongst which three analyzed the solid fraction, and three analyzed the supernatant/filtrate fraction.

**Table 3.** Regression coefficients for individual studies correlating SARS-CoV-2 measurements in wastewater (copies / mL) with COVID-19 case data of associated locations.

**Daily new COVID-19 cases per 100,000 inhabitants**
| Author | Linear regresion |  |  |
| --- | --- | --- | --- |
|  | Slope | R-squared | p-value |
| D'Aoust, P.M et al. | 0.52 | 0.511 | 1.03E-07 |
| Graham, K et al. | 196.64 | 0.351 | 3.99E-17 |
| Scherchan, S.P et al | 0.03 | 0.169 | 1.44E-01 |
| Peccia, J et al. | 1994.77 | 0.163 | 2.79E-10 |
| Hata, A et al. | 0.16 | 0.0494 | 3.85E-02 |
| Miyani, B et al. | -0.20 | 0.0141 | 5.10E-01 |
| Haramoto, E et al | -4.14 | 0.00358 | 7.29E-01 |
| Trottier, J et al | 0.19 | 0.00294 | 8.54E-01 |
| Nemudryi, A et al. | -0.02 | 0.00282 | 7.65E-01 |

**Cumulative COVID-19 cases per 100,000 inhabitants**
| Author | Linear regresion |  |  |
| --- | --- | --- | --- |
|  | Slope | R-squared | p-value |
| Gonzalez, R et al. | 0.04 | 0.613 | 4.81E-124 |
| Medema, G et al | 14.82 | 0.398 | 1.30E-09 |
| Sherchan, S.P et al. | 0.00 | 0.0938 | 2.87E-01 |
| Haramoto, E et al. | -4.62 | 0.0784 | 9.81E-02 |
| Hata, A et al. | 0.08 | 0.0283 | 1.19E-01 |
| Westhaus, S et al. | -0.01 | 0.0121 | 6.63E-01 |
| Randazzo, W et al (b) | 0.45 | 0.00775 | 5.79E-01 |

**Active COVID-19 cases per 100,000 inhabitants**
| Author | Linear regresion |  |  |
| --- | --- | --- | --- |
|  | Slope | R-squared | p-value |
| D'Aoust, P.M et al. | 3.85 | 0.329 | 4.70E-05 |
| Randazzo, W et al (a) | 1.61 | 0.107 | 1.19E-01 |
| Westhaus, S et al. | -0.01 | 0.00774 | 7.28E-01 |

Methodological variability was present in all steps of sample collection and analysis procedures (Figure 2). In terms of sampling locations within a wastewater system, most studies analyzed samples collected at the WWTP (16 studies) (D’Aoust, Mercier, et al., 2021; Medema, Heijnen, et al., 2020; Peccia, Zulli, et al., 2020; Kumar, Patel, et al., 2020; Gonzalez, Curtis, et al., 2020; Graham, Loeb, et al., 2020; D’Aoust, Mercier, et al., 2021; Hata, Hara-Yamamura, et al., 2021; Nemudryi, Nemudraia, et al., 2020; Saguti, Magnil, et al., 2021; Sherchan, Shahin, et al., 2020; Trottier, Darques, et al., 2020; Randazzo, Truchado, et al., 2020; Randazzo, Cuevas-Ferrando, et al., 2020; Ahmed, Bertsch, Bivins, et al., 2020; Westhaus, Weber, et al., 2021; Haramoto, Malla, et al., 2020). A much smaller numbers of studies sampled at locations in the sewage collection network (two studies) (Baldovin, Amoruso, et al., 2021; Miyani, Fonoll, et al., 2020) or in-premise (one study) (Baldovin, Amoruso, et al., 2021). Kitamura, K et al. examined SARS-CoV-2 viruses in wastewater at both municipal sewage network locations and WWTP influent samples (Kitamura, Sadamasu, et al., 2021). Among the studies sampling at WWTPs, the service population ranged from 12,770 to 3.2 million individuals, and covered territories in the Americas, Asia, Europe, and Oceania.

**Figure 2.**
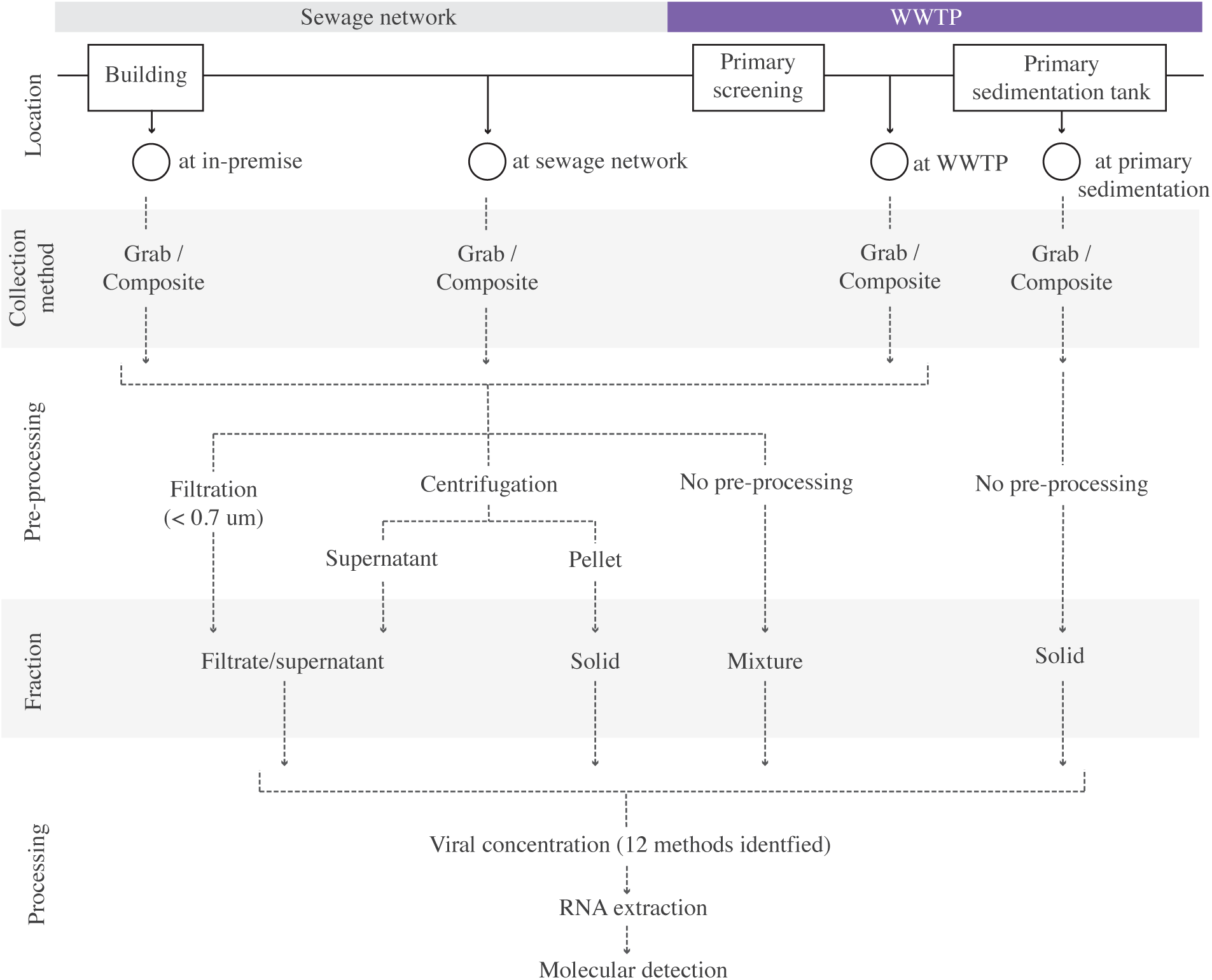
Diagram depicting reported sample collection locations, pre-processing methodologies, and their respective annotations as sampling locations and fractions in this study.

Upon sample collection, studies showed great variability in sample pre-processing conditions, resulting in the enrichment of different wastewater fractions (Figure 2). Supernatant/filtrate fractions were recovered in 12 studies by means of centrifugation between 1.840 and 10.000 g (Medema, Heijnen, et al., 2020; Kumar, Patel, et al., 2020; Gonzalez, Curtis, et al., 2020; Graham, Loeb, et al., 2020; Hata, Hara-Yamamura, et al., 2021; Kitamura, Sadamasu, et al., 2021; Nemudryi, Nemudraia, et al., 2020; Saguti, Magnil, et al., 2021; Sherchan, Shahin, et al., 2020; Trottier, Darques, et al., 2020; Ahmed, Bertsch, Bivins, et al., 2020; Westhaus, Weber, et al., 2021), while two studies retrieved this fraction by filtrating raw wastewater through 0.22 (Baldovin, Amoruso, et al., 2021) and 0.7 µm membranes (Gonçalves, Koritnik, et al., 2021) respectively. Mixed supernatant and suspended solid fractions were identified in six studies where liquid wastewater samples were not subjected to any type of preprocessing. Solid fractions were retrieved in one study from influent wastewater by pellet collection after centrifugation at 1840 g (Kitamura, Sadamasu, et al., 2021), while the remaining three studies utilizing solid fractions collected sludge samples directly from primary sedimentation tanks (Peccia, Zulli, et al., 2020; Graham, Loeb, et al., 2020; D’Aoust, Mercier, et al., 2021). It is important to highlight that a study may pre-process for more than one fraction (Table 2).

Once a fraction of choice was generated, a viral concentration step was usually performed prior to RNA extraction. The viral concentration protocols relied on principles of molecular weight cutoff achieved through ultrafiltration at 10.000 kDa (Medema, Heijnen, et al., 2020; Baldovin, Amoruso, et al., 2021; Gonçalves, Koritnik, et al., 2021; Kitamura, Sadamasu, et al., 2021; Nemudryi, Nemudraia, et al., 2020; Sherchan, Shahin, et al., 2020; Trottier, Darques, et al., 2020; Ahmed, Bertsch, Bivins, et al., 2020; Westhaus, Weber, et al., 2021), the affinity of enveloped viruses to electro-negative membranes, electro-positive membranes, or other adsorbants/flocculants such as PEG, skimmed milk, or aluminum (Kumar, Patel, et al., 2020; Gonzalez, Curtis, et al., 2020; Graham, Loeb, et al., 2020; D’Aoust, Mercier, et al., 2021; Hata, Hara-Yamamura, et al., 2021; Kitamura, Sadamasu, et al., 2021; Sherchan, Shahin, et al., 2020; Randazzo, Truchado, et al., 2020; Randazzo, Cuevas-Ferrando, et al., 2020; Ahmed, Bertsch, Bivins, et al., 2020; Miyani, Fonoll, et al., 2020; Haramoto, Malla, et al., 2020), or a combination of both mechanisms sequentially (D’Aoust, Mercier, et al., 2021; Saguti, Magnil, et al., 2021; Haramoto, Malla, et al., 2020). Some protocols did not include a concentration step and performed RNA extraction directly on the solid fraction (Peccia, Zulli, et al., 2020; Graham, Loeb, et al., 2020). The methodological choices in the concentration step are highly variable, and the twelve different workflows were reported. Reviews on the viral concentration methodology and method evaluation employing surrogates can be found elsewhere (Ahmed, Bertsch, Bivins, et al., 2020; Barril, Pianciola, et al., 2021; Philo, Keim, et al., 2021; Silverman and Boehm, 2020; La Rosa, Bonadonna, et al., 2020; Bofill-Mas and Rusiñol, 2020).

Notably, the various choices in separation methods result from an underlying assumption of differential enrichment/partitioning of the viral particles within the fractions in a sample. Therefore, we considered the fractions as subgroups in achieving pooled estimates of SARS-CoV-2 titers in wastewater (3.2).

### 3.2 Meta-analysis on the positivity of SARS-CoV-2 positive detections from untreated wastewater

While all the current studies took place during the pandemic, the detections of SARS-CoV-2 from wastewater were not always positive. We first ask, what was the grand mean of positivity of detection among studies taking place in the first year of wastewater-based SARS-CoV-2 monitoring? We examined the overall positivity across 1,877 observations in 20 studies, which was 0.67 and a 95%-CI [0.56, 0.79]. Because the sampling mode (i.e., grab or composite sampling) and fractions for analysis (i.e., supernatants/filtrates, mixed supernatant and suspended solids, and solids-only) were expected to introduce variations, we examined the means of the proportion of positive detection by sampling modes (Figure 3) or fractions for analysis (Figure 4). Wastewater SARS-CoV-2 measurements in composite sampling mode had a detection rate of 0.70 and a 95%-CI of [0.47; 0.94], whereas those generated from the grab sampling had an average detection rate of 0.56 and a 95%-CI of [0.32; 0.79]. The SARS-CoV-2 viral detection from the composite sampling approach was significantly higher than those from the grab sample mode (one-sided t-test, p_BH-adjusted_=5.63×10^−9^). When grouped by fractions for analysis, the supernatant, mixed supernatant and suspended solids, and solid fractions exhibited positive proportions of 0.53 (95%-CI [0.32; 0.75]), 0.62 (95%-CI [0.12; 1]), and 0.82 (95%-CI [0.43; 1]), respectively. Solid and solid-supernatant mixtures had significantly higher average positive proportion than the supernatant/filtrate fraction (p_BH-adjusted_<2×10^−16^ and p_BH-adjusted_=2.60×10^−10^ in pairwise t-test, respectively). Solid analysis exhibited a significantly higher average positive proportion than the solid-supernatant mixtures (p_BH-adjusted_=6.50×10^−8^). It should be noted that even within subgroups, high heterogeneity (I-squared 0.97-0.99) was revealed from the metanalysis. This could be caused by the variations in COVID-19 cases or other local variables associated with a study, which will be explored in the regression analysis in Section 3.4.

**Figure 3.**
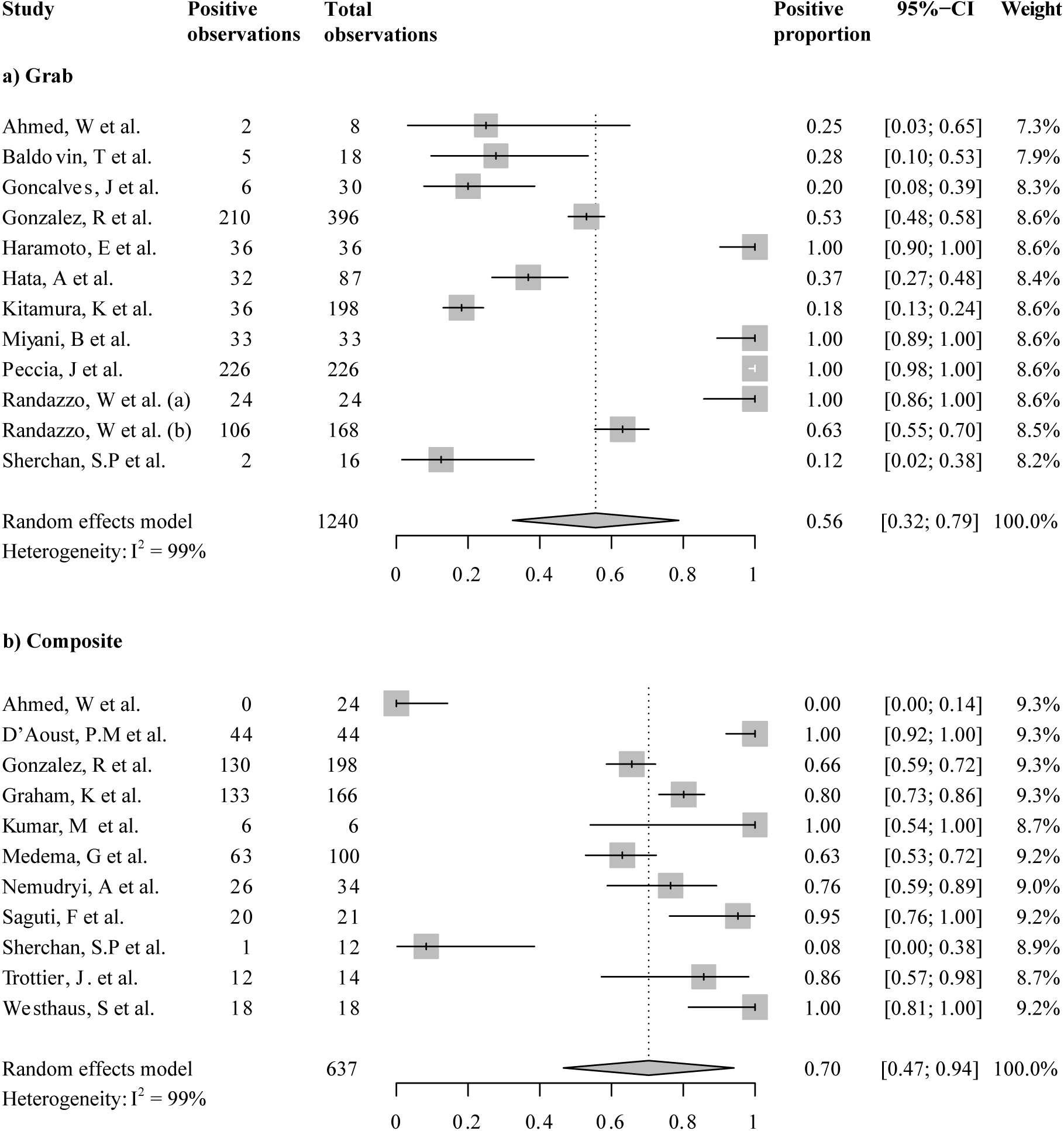
Forest plot of selected aggregation reporting the proportions of positive detections for SARS-CoV-2 in wastewater samples. Pooled estimates for (a) all studies utilizing grab samples and (b) all studies utilizing composite samples.

**Figure 4.**
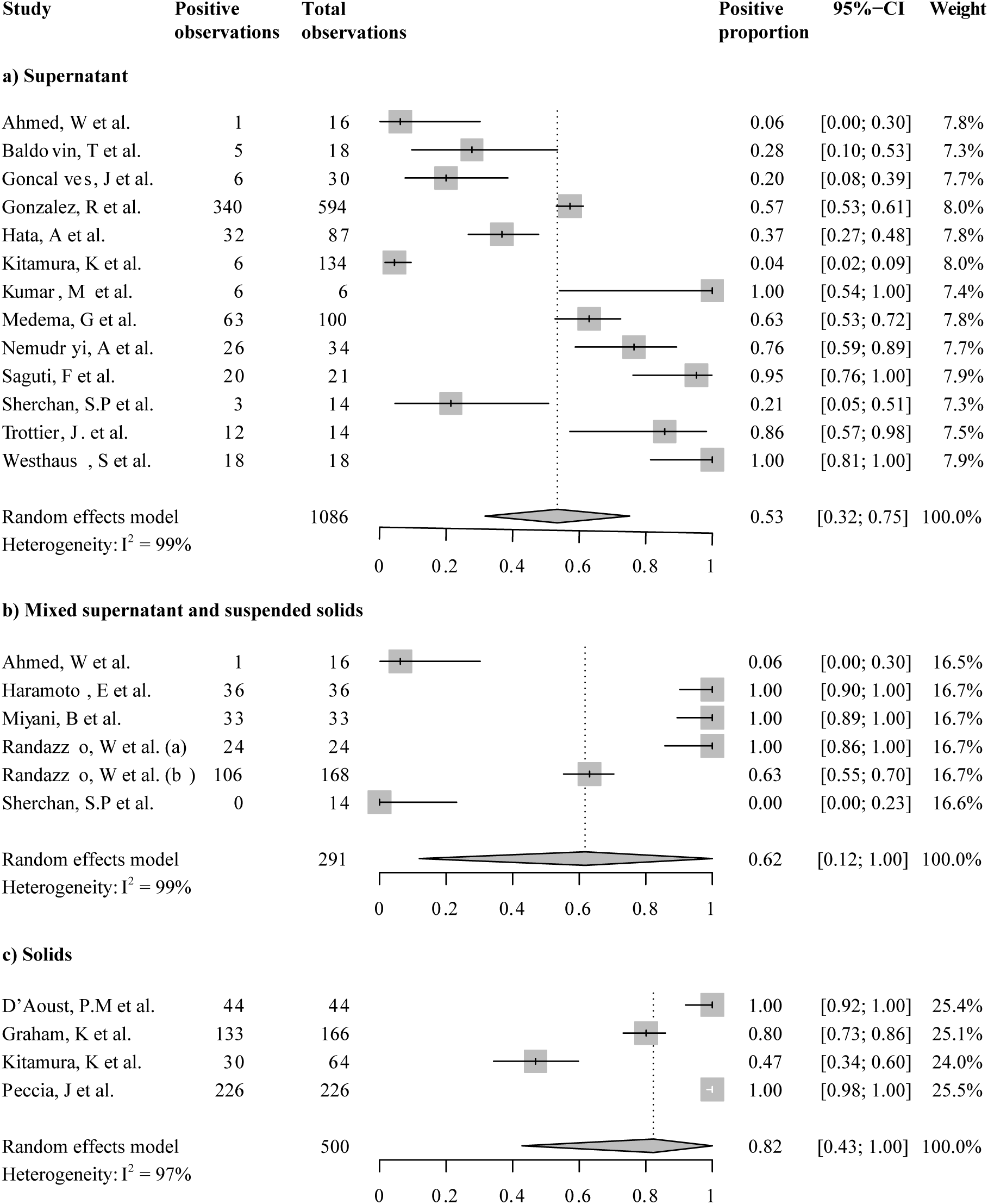
Forest plot of selected aggregation reporting the proportions of positive detections for SARS-CoV-2 in wastewater samples. Pooled estimates for (a) all studies analyzing supernatant/filtrates (b) all studies analyzing mixtures without pre-processing, and (c) all studies analyzing solids.

### 3.3 Meta-analysis of SARS-CoV-2 titers in untreated wastewater

We focused on studies that reported SARS-CoV-2 titers as units per volume to calculate a pooled estimate of SARS-CoV-2 titers in wastewater. These are seventeen studies including a total of 1,674 observations. Across these studies, the average SARS-CoV-2 titers was 5,244.37 copies/mL (95%-CI [0; 16,432.65])

We then aggregated studies by the fractions analyzed, i.e., supernatants/filtrates, mixed supernatant and suspended solids, and solids. A forest plot showing the study means, weighted subgroup means, and confidence intervals is displayed in Figure 5. The average titers in wastewater supernatant, mixture, and solids are 49.83 copies/mL (95%-CI [0; 136.87]), 180.7 copies/mL (95%-CI [0; 510.73]), and 30,455.7 copies/mL (95%-CI [0; 161,832.78]), respectively. Viral titers from solid analysis exhibited significantly higher means than the other two groups (p_BH-adjusted_<2e-16 for both comparisons), yet the other two groups did not significantly differ (p_BH-adjusted_ >0.97). This finding suggests that once the viral titers were beyond detection limits, the difference between analyzing supernatants/filtrates or the mixture was not as strong.

**Figure 5.**
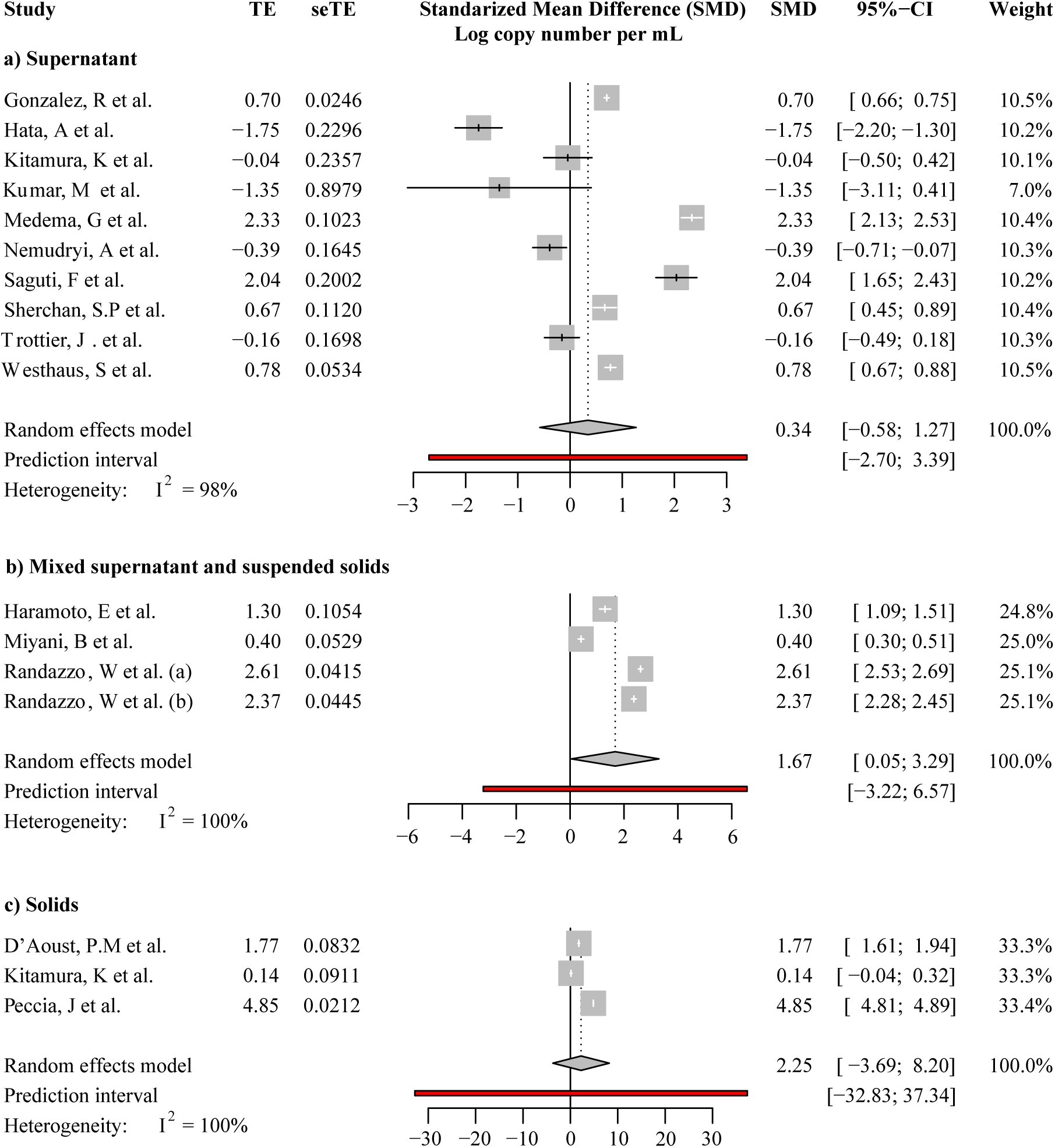
Forest plot of selected aggregations reporting weighted means of SARS-CoV-2 titers in wastewater. Pooled estimates for (a) all studies analyzing supernatant/filtrates (b) all studies analyzing mixtures without pre-processing, and (c) all studies analyzing solids. The forest plot includes data from all studies that reported SARS-CoV-2 titers as copy numbers per unit volume. CI-confidence interval.

Notably, viral titers varied largely even among studies investigating SARS-CoV-2 titers in the same sample fractions, as shown by heterogeneity across studies (*I*^*2*^) higher than 95% in all subgroups (Figure 5). We further aggregated the observations by grab/composite and focused on studies that reported WWTP samples alone (Figure S5). The cross-study heterogeneity remained high (*I*^*2*^ > 93%) even after data were aggregated in such more methodologically homogenous groups. The observed heterogeneity suggested that pandemic severity, as well as other local variables, may drive the variations in SARS-CoV-2 titers among studies.

### 3.4 Correlation between SARS-CoV-2 titers in wastewater and reported COVID-19 cases

The overall correlation between daily new cases and SARS-CoV-2 titers is 0.28 (95%-CI, [0.01; 0.51, Table S3). The Pearson Rho between cumulative case and SARS-CoV-2 titers as measured by Pearson Rho was 0.29 (95%-CI, [-0.15; 0.73], Table S3). For both kinds of epidemiological indicators, wastewater-based SARS-CoV-2 viral titers exhibited overall positive correlation.

### 3.5 What drive the variations in SARS-CoV-2 titers in wastewater?

We ask, how much of the large heterogeneity in the average copy numbers of SARS-CoV-2 in wastewater can be explained by sampling mode, fraction for analysis, and COVID-19 case prevalence, respectively? To answer this question, we built univariate models focusing on each covariate, respectively. Studies reporting cumulative cases (N_[observations]_ = 912, N_[studies]_ = 8) and daily cases (N_[observations]_ = 500, N_[studies]_ = 8) were examined separately to ensure consistent within-group case reporting units.

First, we built logistic regression models to explain the relationships between positive SARS-CoV-2 detection from sewage and each covariate considered. The models with sampling mode and fraction of analysis as sole predictors explained 0.6% and 12.4% (Tjur’s R-squared) of the total variability in SARS-CoV-2 positive detections, respectively (Table 4). The proportion of variances explained by daily and cumulative cases were 9.3% and 5.9%, respectively (Table 5).

**Table 4.** Methodological variables explaining variances in SARS-CoV-2 positive detections and titers.

| Univariate models | Binomial (N<sub>obs</sub> = 1508) |  |  | Gaussian (log transformation on titers) (N<sub>obs</sub> = 936) |  |  |
| --- | --- | --- | --- | --- | --- | --- |
|  | Coefficient [95%-CI] | p-values | Explained variance (R-squared) | Coefficient [95%-CI] | p-values | Explained variance (R-squared) |
| Grab_composite |  |  | 0.006 |  |  | 0.069 |
| Intercept | 0.7480 [0.5505 0.9502] | 2.1 x 10<sup>-13</sup> *** |  | 2.4902 [1.9952 2.9852] | < 2 x 10<sup>-16</sup> *** |  |
| Grab_composite: Grab | -0.3565 [-0.5925 -0.1239] | 0.00284 ** |  | 2.545 [1.9447 3.1456] | 3.1 x 10<sup>-16</sup> *** |  |
| Fraction |  |  | 0.124 |  |  | 0.560 |
| Intercept | 0.0099 [-0.1135 0.1334] | 0.875 |  | 1.5115 [1.2497 1.7733] | < 2 x 10<sup>-16</sup> *** |  |
| Fraction:Solid | 2.1675 [1.8058 2.5591] | < 2 x 10<sup>-16</sup> *** |  | 7.5277 [7.0983 7.9570] | < 2 x 10<sup>-16</sup> *** |  |
| Fraction:Solid-supernatant mixture | 1.2664 [0.8874 1.6685] | 1.9 x 10<sup>-10</sup> *** |  | 2.1430 [1.5617 2.7243] | 9.7 x 10<sup>-13</sup> *** |  |

**Table 5.** Fixed-effects and mixed-effects modeling of the effects of COVID-19 daily new cases or cumulative cases on the positive detection and titers of SARS-CoV-2 viruses in wastewater. CI: confidence interval, AIC: Akaike information criterion, BIC: Bayesian information criterion, Loglik: Log likelihood.

| Model name | Daily case model (N<sub>[observations]</sub> = 500, N<sub>[studies]</sub> = 8) |  |  |  | Cumulative case model (N<sub>[observations]</sub> = 912, N<sub>[studies]</sub> = 8) |  |  |  |
| --- | --- | --- | --- | --- | --- | --- | --- | --- |
|  | Binomial |  | Gaussian<br>(log transformation on titers) |  | Binomial |  | Gaussian<br>(log transformation on titers) |  |
|  | Fixed effects | Mixed effects | Fixed effects | Mixed effects | Fixed effects | Mixed effects | Fixed effects | Mixed effects |
| **Fixed effects** | b<br>[95% CI] | b<br>[95% CI] | b<br>[95% CI] | b<br>[95% CI] | b<br>[95% CI] | b<br>[95% CI] | b<br>[95% CI] | b<br>[95% CI] |
| Intercept | 0.6494<br>[0.3341, 0.9646] | 3.3253<br>[-1.3492, 7.9997] | 6.2329<br>[5.6278, 6.8380] | 1.7822<br>[-1.0955, 4.6598] | -0.0350<br>[-0.1976, 0.1265] | -0.0637<br>[-3.9539, 3.8265] | 1.7848<br>[1.5272, 2.0425] | 1.0549<br>[-1.2556, 3.3654] |
| Daily new cases | 0.0630<br>[0.0424, 0.0865] | 0.1064<br>[0.04792, 0.1649] | 0.0107<br>[0.0011, 0.0202] | 0.0061<br>[0.0035, 0.0086] | -- | -- | -- | -- |
| Cumulative cases | -- | -- | -- | -- | 0.0018<br>[0.0012, 0.0025] | 0.0054<br>[0.0041, 0.0066] | 0.0007<br>[0.0001, 0.0013] | 0.0017<br>[0.0013, 0.0020] |
| **Random effects** |  |  |  |  |  |  |  |  |
| Study_ID (variance) | -- | 23.15 | -- | 17.124 | -- | 29.75 | -- | 10.926 |
| Adjusted R<sup>2</sup> | 0.0935 | -- | 0.0093 | -- | 0.0592 | -- | 0.0084 | -- |
| Marginal R<sup>2</sup> | -- | 0.5399 | -- | 0.0056 | -- | 0.0832 | -- | 0.0239 |
| Conditional R<sup>2</sup> | -- | 0.9427 | -- | 0.9253 | -- | 0.9087 | -- | 0.8992 |
| AIC | 422.2 | 200.3590 | 2579.109 | 1406.1 | 1213.9 | 1012.1423 | 2381.286 | 1691.9 |
| BIC | 430.6259 | 213.0028 | 2591.157 | 1422.2 | 1223.536 | 1026.5892 | 2394.007 | 1708.9 |
| Loglik | -209.098 | -97.179 | -1286.55 | -699.07 | -604.95 | -503.07 | -1187.64 | -841.95 |
| Random effects (p-values) | -- | < 2.2 x 10<sup>-16</sup> *** | -- | < 2.2 x 10<sup>-16</sup> *** | -- | < 2.2 x 10<sup>-16</sup> *** | -- | < 2.2 x 10<sup>-16</sup> *** |

Next, we built linear models to examine the relationships between logarithmic transformed viral titers and each covariate. The variance in titers explained by sampling mode and fraction of analysis were 6.9% and a notable 56.0%, respectively, whereas the variance explained by daily and cumulative cases were 0.9% and 0.8%, respectively. In all these models, the role of methodological variables and epidemiological indicators were significant (p<0.005, Table 4 and 5). Daily or cumulative cases and sampling mode explained comparable proportions of variances. Notably, fraction of analysis explained dramatically higher variance in titers than any other variables.

### 3.6 Slope coefficients in Generalized linear models

Successful detection of the virus from wastewater is fundamental to WBE; our generalized linear models on positive detections can provide quantitative insights on the magnitude at which changes in each variable increase the chance of the positive detections (Table 4 Binomial family models and Table 5 Binomial Fixed-effects model). From our models, the odds of positive detection decrease by a factor of 1.43 (95%-CI [1.81, 1.13]) when utilizing grab sampling in contrast to composite sampling. The odds of positive detection increase by a factor of 8.16 (95%-CI [6.08, 12.92]) from solid fraction in contrast to supernatants/filtrates; they increase by a factor of 3.52 (95%-CI [2.43, 5.30]) from solid-supernatant mixture in contrast to supernatants/filtrates. With an increase in active case prevalence of 10 per 100,000 inhabitants, the odds of positive detection increase by a factor of 1.06 unit (95%-CI [1.04, 1.09]); with an increase in cumulative cases of 10 per 100,000 inhabitants, they increase by a factor of 1.02 (95%-CI [1.01, 1.03]).

### 3.7 Mixed-effects model help account for variation by studies

While many applications of WBE rely on positive correlations between SARS-CoV-2 titers in wastewater and disease prevalence, larger or comparable variability was explained by methodological covariates than the reported case prevalence in our models (Table 4 and 5). While it is a consensus that documenting methodological covariates in WBE studies is crucial, learning about important variables about WBE is an ongoing process. To address the need of building explanatory or predictive models in WBE, we considered a mixed-effects framework for modeling SARS-CoV-2 viral titers from multiple studies. Here, we treat studies as a collective source of variance. We hypothesize that in addition to the role of cases as a source of fixed effects on the wastewater measurements, each study presents a source of a study-specific intercept. We tested for the significance of random effects. For both, positivity or titers from daily or cumulative cases, the random effects from the studies were significant (*p-value* < 2.2×10^−16^, Table 5). Mixed effects models also showed a lower AIC or BIC than the corresponding fixed-effects models, suggesting better fits to the data.

For a mixed effects model, we examined both the marginal R-squared, which is the proportion of variance explained by the fixed effects alone (case daily or case cumulative), and the conditional R-squared, which describes the proportion of variance explained by both the fixed and random factors (cases and the study identities respectively). Notably, mixed models exhibited conditional R-squared close to or over 0.9 for both positivity and titers models reporting daily new cases or cumulative cases (Table 5). Thus, simultaneously considering variability across studies greatly improved our ability to explain the variation in wastewater SARS-CoV-2 measurements.

## 4. DISCUSSION

We synthesized the available evidence on SARS-CoV-2 detection and titers in wastewater during the first year of the COVID-19 pandemic. The combined detection rate across studies was 67% (95%-CI, [0.56; 0.79]). The overall SARS-CoV-2 titers across all processing methods was 5,244.37 copies/mL (95%-CI, [0; 16,432.65]). The overall correlation between SARS-CoV-2 titers in wastewater and daily new cases was 0.28 [0.01; 0.51], and the correlation between titers and cumulative cases was 0.29 [-0.15; 0.73]. The overall positive associations reinforce that wastewater is a favorable data source to track COVID-19 dynamics in a community backed by meta-analysis.

Of interest, sampling modes and wastewater fractions had strong influences on the pooled means in proportions of positive detection and SARS-CoV-2 titers. Composite sampling mode had a higher detection rate than grab sampling, as seen from an average detection rate of 0.70 [0.47; 0.94]) and 0.56 (95%-CI, [0.32; 0.79], Figure 3), respectively. Supernant/filtrates, solid-supernatant mixture, and solid fractions increased by average detection rates 0.53 (95%-CI [0.32; 0.75], 0.62 (95%-CI [0.12; 1]), and 0.82 (95%-CI [0.43; 1]), respectively, (Figure 4). Sampling mode and fraction of analysis explained 0.6% and 12.4% of the variance in the proportions of positive detection, and fraction of analysis explained 56% of variance in titers. These results can be useful in designing experimental workflows for WBE. In particular, the large variance in titers explained by fraction of analysis and the large magnitudes in regression coefficients suggest that standardizing the fraction of analysis need to be prioritized when researchers would like to design monitoring efforts across multiple labs. The overall detection rate and those in subgroups of any sewage fraction was below one, suggesting a need for tools to maximize the chance of SARS-CoV-2 detection from sewage samples.

In our meta-analysis, large heterogeneity was detected in all effect sizes investigated (i.e., proportions of positive detections, titers, and Pearson’s Rhos between titers and daily or cumulative cases, Figures 3, 4, and 5, Table S3). We hypothesize that the unexplained variations in SARS-CoV-2 titers detected in wastewater can be affected by study-level factors specific to the wastewater collection system, individual-based testing efforts, and methodological choices. In our meta-analysis, we found that metadata about the collection system, such as per capita water consumption, relative contributions of domestic vs. commercial/industrial water, or sewage travel times (i.e., residence times) are currently rare. These collection system-level variables can affect the dilution of fecal materials and the genetic decay of the viral signal (Silverman and Boehm, 2020; Foladori, Cutrupi, et al., 2020; Ahmed, Bertsch, Bibby, et al., 2020). Thus, more detailed metadata reporting regarding the wastewater collection system is needed to better explain variations across sites. Lately, it was proposed that wastewater be viewed as an independent indicator of true prevalence, as epidemiological indicators from current reporting can be affected by under-reporting (Olesen, Imakaev, et al., 2021). Therefore, methods and tools to interrogate the wastewater metagenome and derive system-level data, or bridging wastewater-based measurements to prevalence, deserve more attention.

It should be noted that heterogeneity in titers and correlations observed here may not be fully explained by recovery efficiencies of viruses from wastewater samples during viral concentration workflows. To illustrate this complexity, we discuss two studies where recovery efficiencies were reported. In one study, an average viral titer of 881 ± 633 copies/mL was detected when COVID-19 in the associated area was between 10-80 cumulative cases per 100,000 inhabitants (Medema, Heijnen, et al., 2020); in another study, an average viral titer of 1.9 ± 6.0 marker copies/mL was reported with the same range of COVID-19 prevalence (10-80 cumulative cases per 100,000 inhabitants). After adjusting titers by reported recovery efficiencies (73 and 7.7%, respectively), the adjusted copy numbers (1,206 and 27 marker copies/mL, respectively) still vary by two orders of magnitude.

While the field’s ability to quantify the effects from methodological variables and collection systems are important ongoing research topics, mixed-effects models treating “studies” as a source of random effects can be considered a useful way to perform inference and prediction. Mixed-effects models handle a wide range of scenarios where observations have been sampled in hierarchical structure rather than completely independently. In this study, treating studies as a source of random effects on intercepts profoundly improved the quality of the model, as seen in improved AIC and BIC compared to respective fixed-effects models (Table 5). The final models reached conditional R-squared values above 0.9. The mixed-effects approach provides an alternative for researchers to leverage existing data from studies conducted elsewhere to build models useful for explaining variations in local observations.

## 5. Study limitations

Our study had several limitations. The most notable is the large amount of unexplained heterogeneity in positive detection, SARS-CoV-2 titers, and Pearson correlations across studies. This is likely attributable to variability in methodological differences in SARS-CoV-2 virus measurements, wastewater-system characteristics, and ways that epidemiological data were collected and reported. Thus, we employed mixed-effects models to make inferences about the correlation between epidemiological indicators and viral detection/titers, treating study-level variations as a source of random effects. Second, the studies resulting from the screening method were primarily carried out at wastewater treatment plants. Future meta-analysis focusing on collection systems or buildings may become possible when more data become available.

## Data Availability

All data produced in the present study are available upon reasonable request to the authors

---

**Figure S1.**
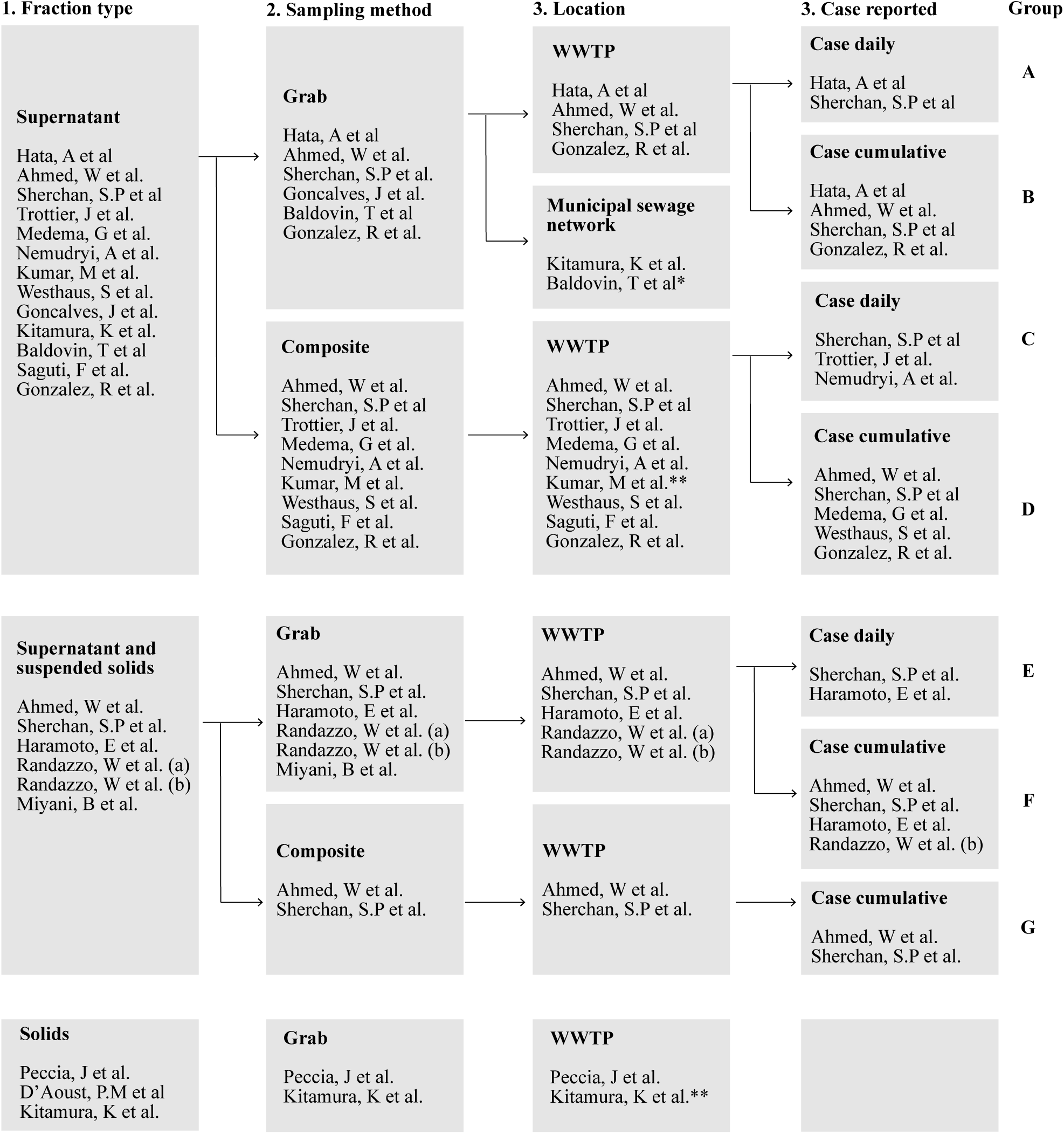
Schematic showing the agregation of studies based i) fraction type, ii) sample collection location and iii) case type reported by study. Two entries are included for Ahmed, W et al. and Kitamura, K et al. as these studies report data for multiple fractions.Studies are listed only if two or more studies fall within the same group. * Baldovin, T et al. and Goncalvez, J et al. reports only qualitative data. ** Cases are reported. However no information was available to allow for normalizatin of daily cases by population size.

**Figure S2.**
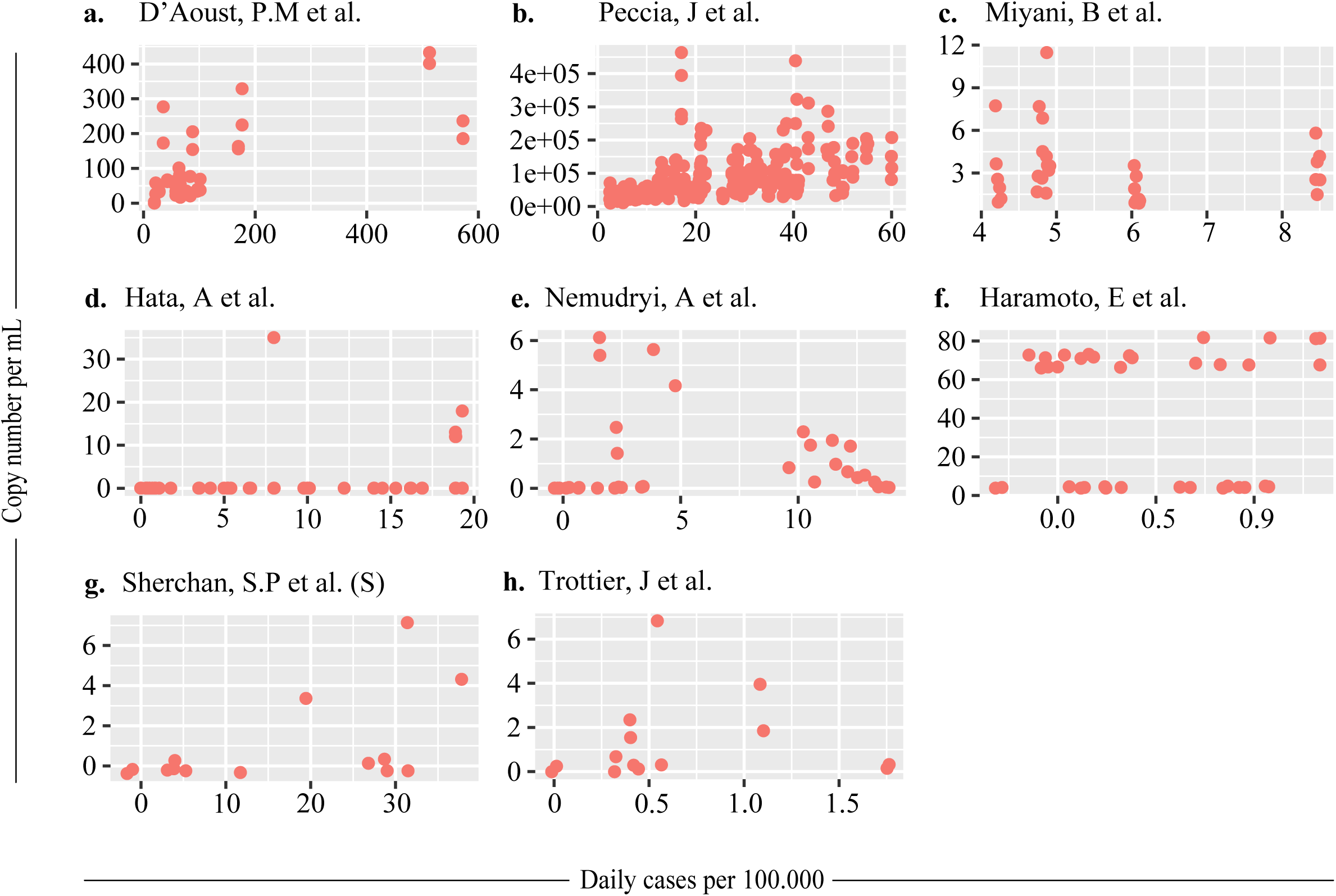
Individual correlations (per study) of SARS-CoV-2 measurements in wastewater with daily COVID cases reported. Daily cases are normalized by population size and reported as cases in 100.000 inhabitants. Only data for supernatant fraction (S) in Sherchan, S.P. et al. is presented, as supernatant and suspended solids fraction did not show positive results for SARS-CoV-2.

**Figure S3.**
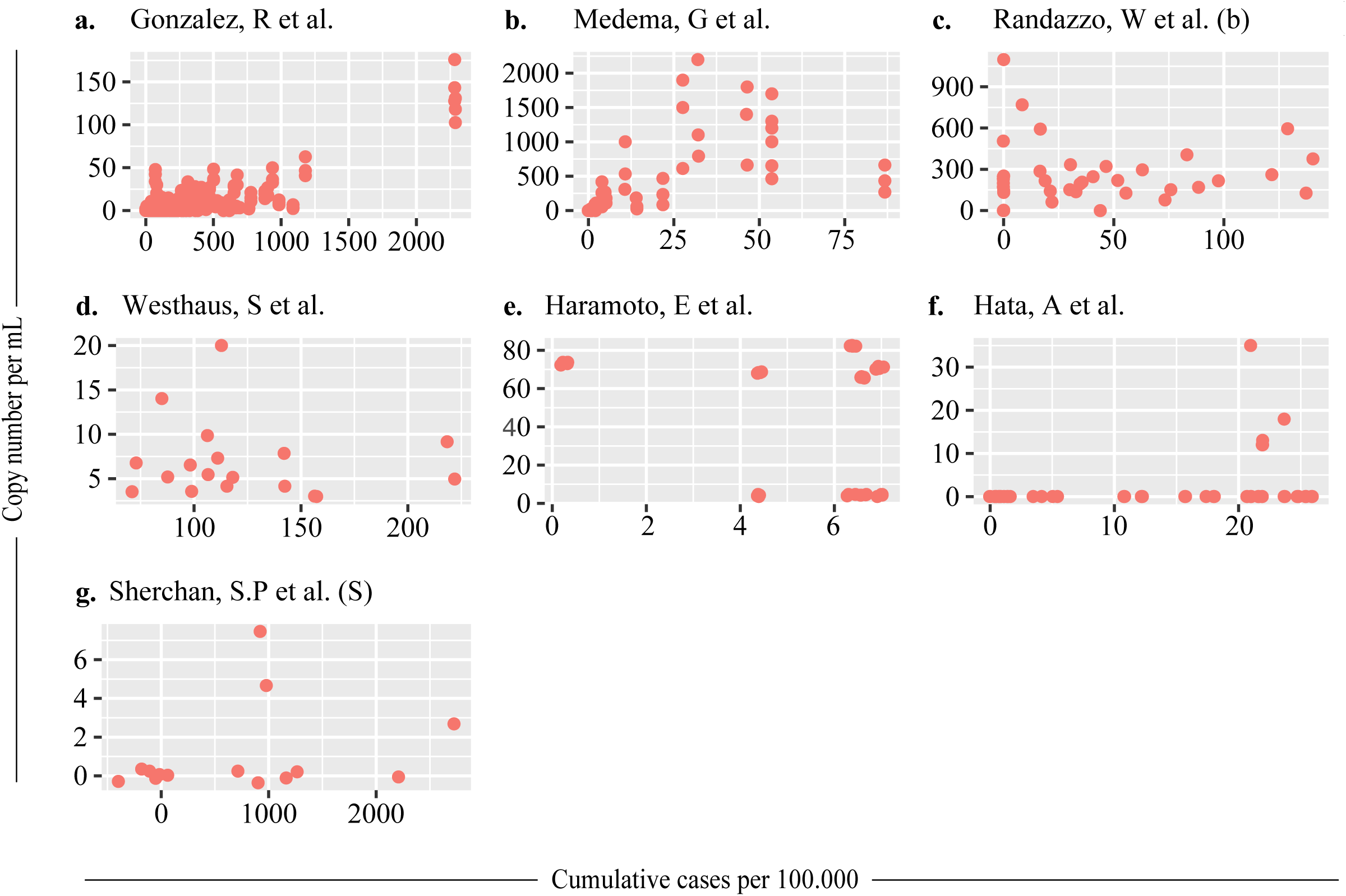
Individual correlations (per study) of SARS-CoV-2 measurements in wastewater with cumulative COVID cases reported. Cumulative cases are normalized by population size and reported as cases in 100.000 inhabitants. Only data for supernatant fraction (S) in Sherchan, S.P. et al. is presented, as supernatant and suspended solids fraction did not show positive results for SARS-CoV-2. Data for Ahmed, W et al. is not presented as only one sample per fraction type was positive for SARS-CoV-2.

**Figure S4.**
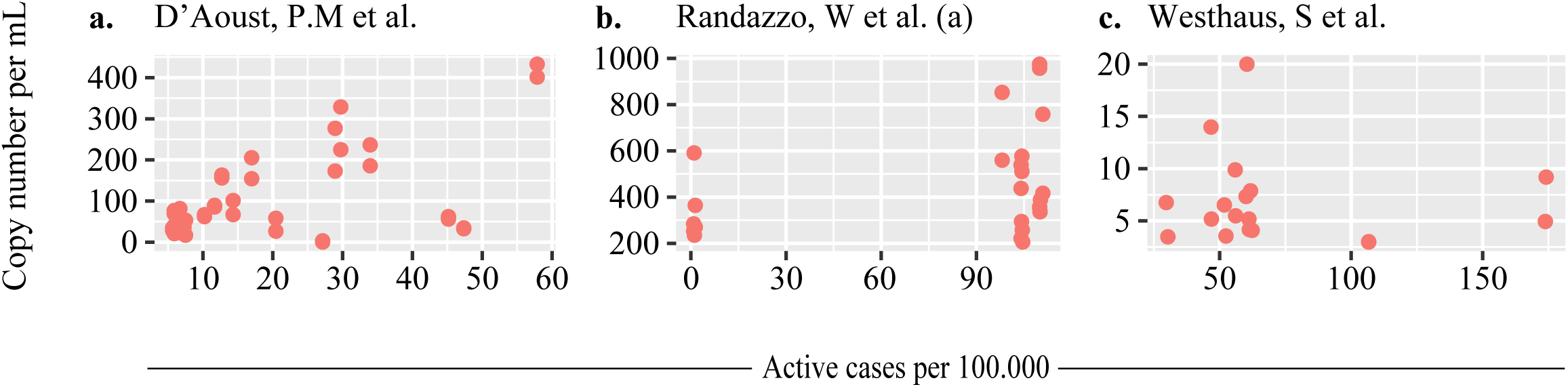
Individual correlations (per study) of SARS-CoV-2 measurements in wastewater with active COVID cases reported. Active cases are normalized by population size and reported as cases in 100.000 inhabitants.

**Figure S5.**
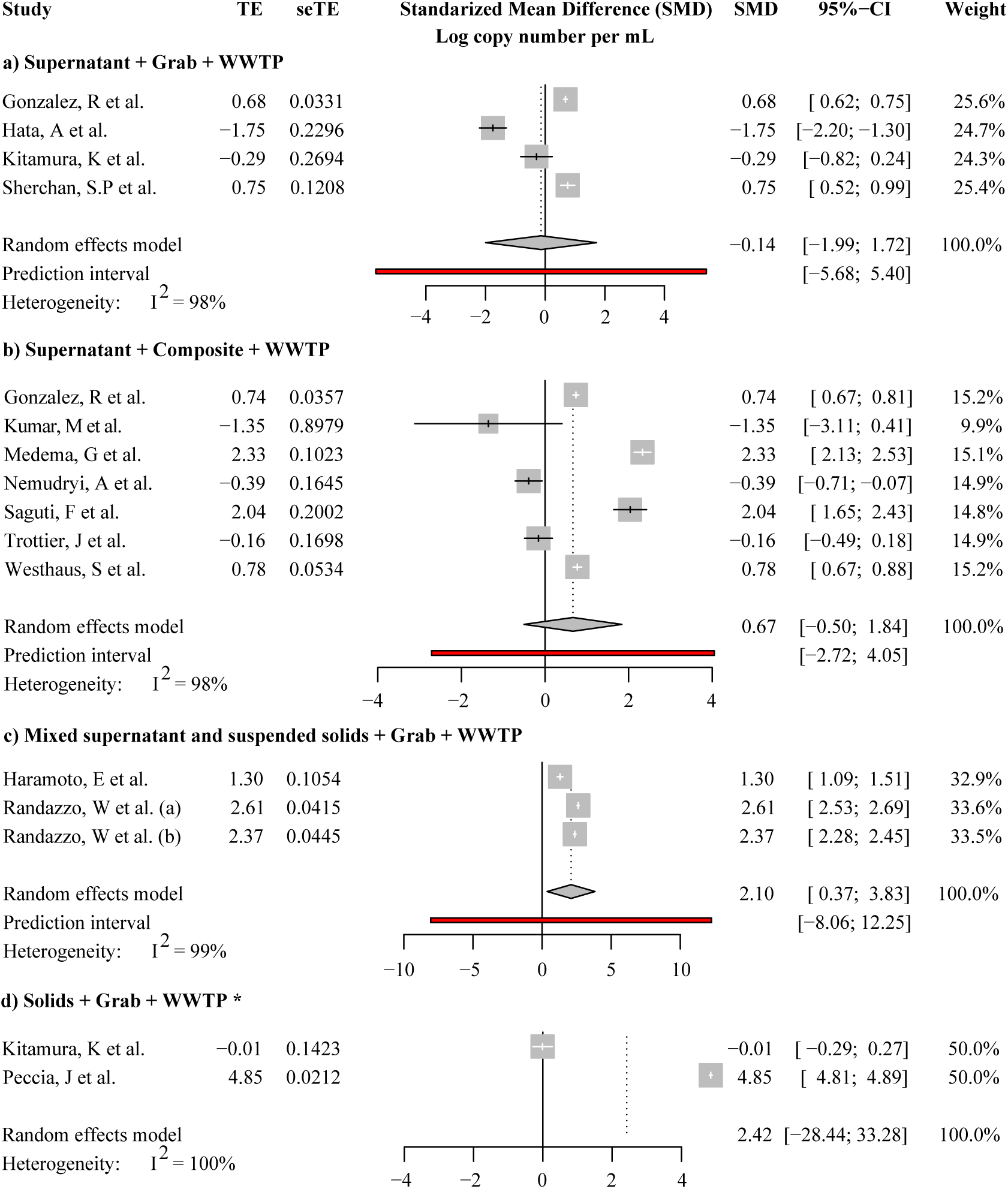
Standardized mean difference of SARS-CoV-2 copy numbers per mL of wastewater. Studies are grouped based on fraction type and location of sample collection. Five studies are not included in the forest plot. These are: i) Graham, K et al. data is not presented as this study uniquely reports SARS-CoV-2 levels per unit of mass, ii and iii) Goncalvez, J et al. and Baldovin, T et al. reports only qualitative data for SARS-CoV-2 measurments in wastewater (quantification cycle for qPCR), and iv and v) Ahmed, W et al. and Sherchan, S.P et al. as theese studies reported only two samples with a poitive result for the detection of SARS-CoV-2 for the respective sample groupings. Kitamura, K et al. examined SARS-CoV-2 levels in both, solids and supernatant and suspended solids samples. Values for each type of sample are presented in the relevant group. Scales between fraction groups differ. Prediction interval can not be calculated for two studies.

